# Cognitive Practice Effects Delay Diagnosis; Implications for Clinical Trials

**DOI:** 10.1101/2020.11.03.20224808

**Authors:** Mark Sanderson-Cimino, Jeremy A. Elman, Xin M. Tu, Alden L. Gross, Matthew S. Panizzon, Daniel E. Gustavson, Mark W. Bondi, Emily C. Edmonds, Graham M.L. Eglit, Joel S. Eppig, Carol E. Franz, Amy J. Jak, Michael J. Lyons, Kelsey R. Thomas, McKenna E. Williams, William S. Kremen, Alzheimer’s Disease Neuroimaging Initiative

## Abstract

**Objective:** Practice effects on cognitive tests obscure decline, thereby delaying detection of mild cognitive impairment (MCI). This reduces opportunities for slowing Alzheimer’s disease progression and can hinder clinical trials. Using a novel method, we assessed the ability of practice-effect-adjusted diagnoses to detect MCI earlier, and tested the validity of these diagnoses based on AD biomarkers.

**Methods:** Of 889 Alzheimer’s Disease Neuroimaging Initiative participants who were cognitively normal (CN) at baseline, 722 returned at 1-year-follow-up (mean age=74.9±6.8). Practice effects were calculated by comparing returnee scores at follow-up to demographically-matched individuals who had only taken the tests once, with an additional adjustment for attrition effects. Practice effects for each test were subtracted from follow-up scores. The lower scores put additional individuals below the impairment threshold for MCI. CSF amyloid-beta, phosphorylated tau, and total tau were measured at baseline and used for criterion validation.

**Results:** Practice-effect-adjusted scores increased MCI incidence by 26% (p<.001). Adjustment increased proportions of amyloid-positive MCI cases (+20%) and reduced proportions of amyloid-positive CNs (−6%) (ps<.007). With the increased MCI base rate, adjustment for practice effects would reduce the sample size needed for detecting significant drug treatment effects by an average of 21%, which we demonstrate would result in multi-million-dollar savings in a clinical trial.

**Interpretation:** Adjusting for practice effects on cognitive testing leads to earlier detection of MCI. When MCI is an outcome, this reduces recruitment needed for clinical trials, study duration, staff and participant burden, and can dramatically lower costs. Importantly, biomarker evidence also indicates improved diagnostic accuracy.

## Introduction

Alzheimer’s Disease (AD) is a leading cause of death in adults over age 65^1^ and an estimated 1 in 85 people will be living with the disease by 2050.^2^ Give the protracted AD prodromal period,^3^ emphasis is now on clinical trials that begin with cognitively normal (CN) individuals who may progress to mild cognitive impairment (MCI).^3-6^ Delayed detection of MCI is essentially misdiagnosis, i.e., labeling someone as CN when they, in fact, have MCI. It impedes identification of meaningful drug effects and may lead to misinterpretation of findings in clinical trials.^7,8^ Detection of MCI as early as possible is thus critical.

Repeat cognitive assessments are necessary for accurately determining transitions from CN to MCI or MCI to dementia. However, repeat assessments are impacted by practice effects (PEs) that mask true decline and compromise diagnostic accuracy.^9,10^ PEs have been found across multiple cognitive domains and test-retest intervals as long as 7 years in older adults.^10,11^ PEs after 3-6 months have been observed in those with MCI and even mild AD.^12,13^

A common view of PEs is that they only occur when scores increase over time.^9,13,14^ However, PEs can exist even when there is overall decline, as they may still cause underestimation of decline (Figure 1).^9,13^ In such situations, failure to account for PEs may delay MCI diagnosis because PEs would inflate scores above diagnostic impairment thresholds.^9,10,15,16^

**Figure 1:**
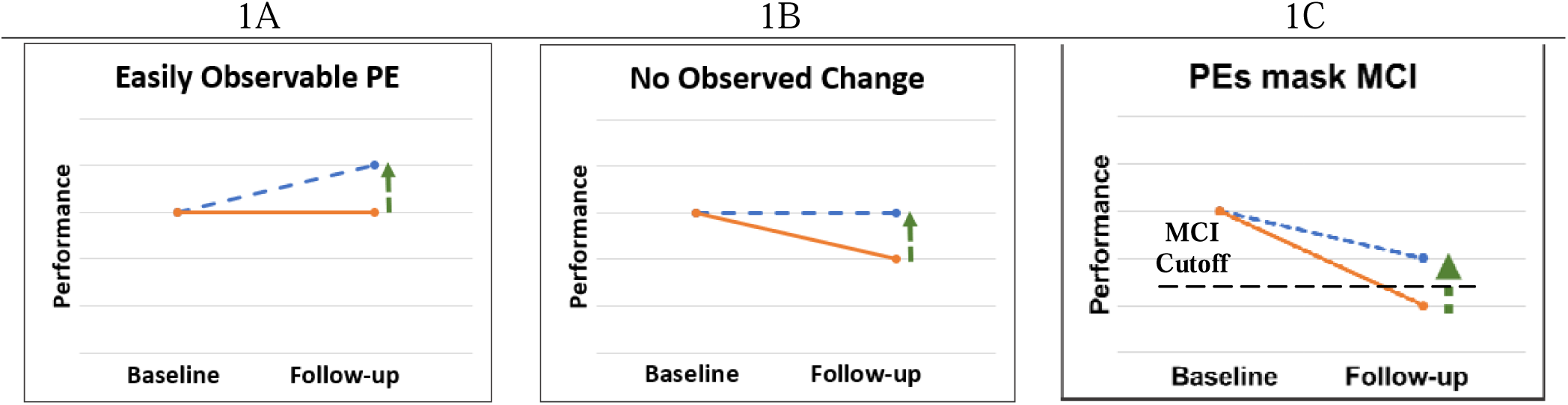
Practice effects with and without true decline. The solid line represents true cognitive ability. The diagonal dashed line represents observed performance, which is inflated due to a practice effect (vertical arrow). **1A**: Typically observed practice effect: an individual’s score increases from baseline to follow-up, demonstrating a typical practice effect. **1B**: Practice effect in the context of cognitive decline. In this scenario, an individual’s score is decreasing overtime. A practice effect still exists but is masked by cognitive decline such that the individual’s performance appears to be stable but is actually better than it would have been without previous exposure to the test. **1C**: Practice effects impair detection of MCI. In this situation, an individual has declined below an MCI cutoff. However, practice effects are inflating their score so that they now fall above the MCI cutoff and will be diagnosed as cognitively normal at follow-up.

Despite their importance, PEs are largely ignored in longitudinal studies, clinical trials, and clinical practice.^9,13,17-19^ A review of PEs in MCI and AD samples noted considerable evidence of PEs (i.e., increased scores) in clinical trials.^13^ However, despite recognition that accounting for PEs may potentially improve clinical trials and diagnostic accuracy, there are minimal empirical data on PEs in clinical trials.^9,13,14^

In a 6-year follow-up, MCI incidence doubled (4.6% vs 9.0%) when scores were adjusted for PEs via a replacement-participants method that is able to gauge PEs even when performance declines.^10^ Increased incidence means earlier detection, but it is crucial to determine if it truly represents more accurate diagnosis rather than methodological artifact since PE adjustment lowers scores.

Here, we employed a novel approach by identifying the equivalent of replacement participants in the Alzheimer’s Disease Neuroimaging Initiative (ADNI). In individuals who were CN at baseline, we hypothesized that: 1) we would observe PEs at the 12-month follow-up; and 2) accounting for PEs would increase the number of MCI diagnoses at follow-up. To demonstrate criterion validity, we hypothesized that: 3) PE-adjusted diagnoses would result in more AD biomarker-positive MCI cases and fewer biomarker-positive CN individuals than PE-unadjusted diagnoses. Finally, we completed power/sample size calculations, hypothesizing that 4) accounting for PEs would substantially reduce the number of participants needed for clinical trials. As a real-world example, we applied these estimates to a hypothetical trial with progression to MCI as a key outcome using recruitment data from a major clinical trial. Earlier and more accurate detection should thus have a substantial impact on clinical trials by reducing study duration, attrition, participant and staff burden, and overall cost.

## Participants and Methods

### Participants

Data used in the preparation of this article were obtained from the ADNI database (adni.loni.usc.edu). The ADNI was launched in 2003 as a public-private partnership, led by Principal Investigator Michael W. Weiner, MD. The primary goal of ADNI has been to test whether serial magnetic resonance imaging, positron emission tomography, other biological markers, and clinical and neuropsychological assessment can be combined to measure the progression of MCI and early AD. For up-to-date information, see www.adni-info.org. Participants from the ADNI-1, ADNI-GO, and ADNI-2 cohorts were included.

We identified 889 individuals who were CN at baseline; 722 of them returned for a 12-month-follow-up. Mean educational level of returnees was 16 years (SD=2.7), 47% were female, and mean baseline age was 74.9 years (SD=6.8). All participants completed neuropsychological testing at baseline and 12-month follow-up. After accounting for PEs, we re-diagnosed returnees at their 12-month follow-up as CN or MCI.

### Procedures

Six cognitive tests were examined across the test-retest interval (mean=12.21 months; SD=0.97). Memory tasks included the Wechsler Memory Scaled-Revised, Logical Memory Story A delayed recall, and the Rey Auditory Verbal Learning Test delayed recall. Language tasks included the Boston Naming Test and Animal Fluency. Attention-executive function tasks were Trails A and Trails B. The American National Adult Reading Test provided an estimate of premorbid IQ. Participants completed the same version of tests at baseline and 12-month visits.

PE-adjusted and unadjusted scores were converted to z-scores based on independent external norms that accounted for age, sex, and education for all tests except the AVLT.^20^ Having found no external norms for the AVLT that were appropriate for this sample and accounted for age, education and sex, the AVLT was z-scored based on the ADNI participants who were CN at baseline (n=889). AVLT demographic corrections were based on a regression model that followed the same approach as the other normative adjustments. Beta values were multiplied by an individual’s corresponding age, education, and sex. The products were then removed from the AVLT raw scores. These adjusted AVLT scores were then z-scored.

We focused primarily on MCI diagnosed according to the Jak-Bondi approach.^7,8,21^ MCI was diagnosed when scores on ≥ 2 tests within the same cognitive domain were each >1 SD below normative means. Compared to MCI classifications using the standard Petersen criteria in ADNI, participants meeting Jak-Bondi criteria for MCI were previously shown to be more likely to progress to dementia and less likely to revert to CN, and demonstrated higher proportions with abnormal biomarkers and *APOE*-*ε*4 alleles.^7,8^ To test whether the results were specific to a particular diagnostic approach, we performed a second set of analyses based on Petersen MCI criteria.^22^

Biomarkers included cerebrospinal fluid amyloid-beta (Aβ), phosphorylated tau (p-tau), and total tau (t-tau) levels collected at baseline. The ADNI biomarker core (University of Pennsylvania) used the fully automated Elecsys immunoassay (Roche Diagnostics). Sample collection and processing have been described previously.^23^ Cutoffs for biomarker positivity were: Aβ+: Aβ<977 pg/mL; p-tau+: p-tau>21.8 pg/mL; t-tau+: t-tau>270 pg/mL (http://adni.loni.usc.edu/methods).^24,25^ There were 521 returnees with Aβ, 518 with p-tau, and 519 with t-tau data.

### Practice effect calculation and statistical analysis

Practice effects were calculated using a modified version of a replacement-participants method.^10^ This method was selected in part because a meta-analysis noted that almost all studies of PEs considered only observed performance increases (Figure 1A), and recommended the replacement-participants methods in situations where decline is expected.^9,15^ A more recent review paper also posited that PEs can still exist when there is a decline in observed performance, but did not cite empirical evidence for that claim.^13^ In some situations PEs will not necessarily manifest as improvements for middle-aged and older adults, particularly for individuals on an AD trajectory.^26^ The replacement-participants approach involves recruiting participants for testing at follow-up who are demographically matched to returnees. The only difference between the groups is that replacements are taking the tests for the first time whereas returnees are retaking the tests. Comparing scores at follow-up between returnees and replacement participants (with additional adjustment for attrition effects) allows for detection of PEs when observed scores remain stable (Figure 1B) and even when they decline (Figure 1C). In both scenarios, scores would have been lower without repeated testing. Thus, the goal of the replacement method is to create follow-up scores over retest intervals that are free of PEs and comparable to general normative data. By design, this method is equally applicable for any sample and any test because the returnees and replacements are always matched on demographic characteristics, test measures, and retest interval.

We are frequently asked how the replacement method compares to other PE methods, especially those that calculate PEs at an individual level. While understandable, in our view these questions are somewhat misguided as the methods address different issues. Studies of very short-term (e.g., 1 week) PEs do not use replacement participants but do typically involve a comparison sample also retested tested after 1 week. Individuals with smaller 1-week PEs— primarily on memory tests—have worse baseline ability, more abnormal baseline biomarkers, and increased risk for future decline and progression to dementia.^14,27-29^ Thus, the short-term PE method is useful for predicting *relative* future decline. However, short-term PEs are not a measure of an individual’s level of functioning at follow-up, especially if the follow-up is after a long retest interval. Short-term PEs are therefore uninformative about when a diagnostic impairment threshold is crossed. In contrast, the replacement method is not intended to predict who is more likely to become impaired, but it can alter when an individual is defined to have crossed a diagnostic threshold at follow-up. It thus has the potential for earlier detection of diagnosis.

Bootstrapping (5,000 resamples, with replacement) was used to calculate PE values for each cognitive test. As shown in Figure 2, the following steps were completed at each iteration of the bootstrap: participants with valid baseline data were identified (n=889). Box 1. Participants who also had 12-month follow-up data comprised the returnees (n=722). Box 2. A random subsample (n=100) of returnees was selected. Box 3 Baseline data for these participants were labeled as Returnees_T1_; follow-up data for these participants were labeled Returnees_T2_. Box 4. The 100 Returnees_T1_ participants were removed from the pool of baseline data, leaving 789 remaining baseline participants. Box 5. From these remaining 789 baseline participants,100 potential pseudo-replacements were selected. This selection was random, except that the potential pseudo-replacements had to be within the age range of the 100 Returnees_T2_ participants. Box 6. We refer to these as pseudo-replacements because ADNI did not recruit matched replacement participants for this purpose. Through an iterative process that added and/or subtracted participants one at time, the potential Pseudo-Replacements_T1_ (N=100) were matched at a mean level to the Returnees_T2_ sample. These groups were constrained to be similar (ps >.70) on comparisons of age, birth sex, education, and premorbid IQ. These factors were chosen because they may affect cognition and/or PE magnitude.^9,10^ Once a mean-matched sample was found, it was labeled Pseudo-Replacments_T1_, and this sample ranged in size from 80 to 120 participants. It did not always equal 100 because a different number of individuals might need to be removed/added at different iterations in order to achieve mean level matching. The Pseudo-Replacments_T1_ sample and the Returnees_T2_ sample were demographically matched and only differed in that the Returnees_T2_ had taken the tests before while Pseudo-Replacmeents_T1_ had taken the tests only once. Box 7. After the—on average—100 Pseudo-Replacments_T1_ were removed from the pool of baseline data, there were an average of 689 remaining baseline participants (Box 4 minus 100). Box 8. 100 potential baseline participants who were age-matched (p >.70) to the returnees at baseline (Returnees_T1_) were selected from that pool. Box 9. After successful matching, this group of participants was labeled Age-Matched Baseline_T1_. The age-matched baseline sample is a subset of participants at baseline who were compared with returnee baseline scores in order to gauge attrition effects.

**Figure 2:**
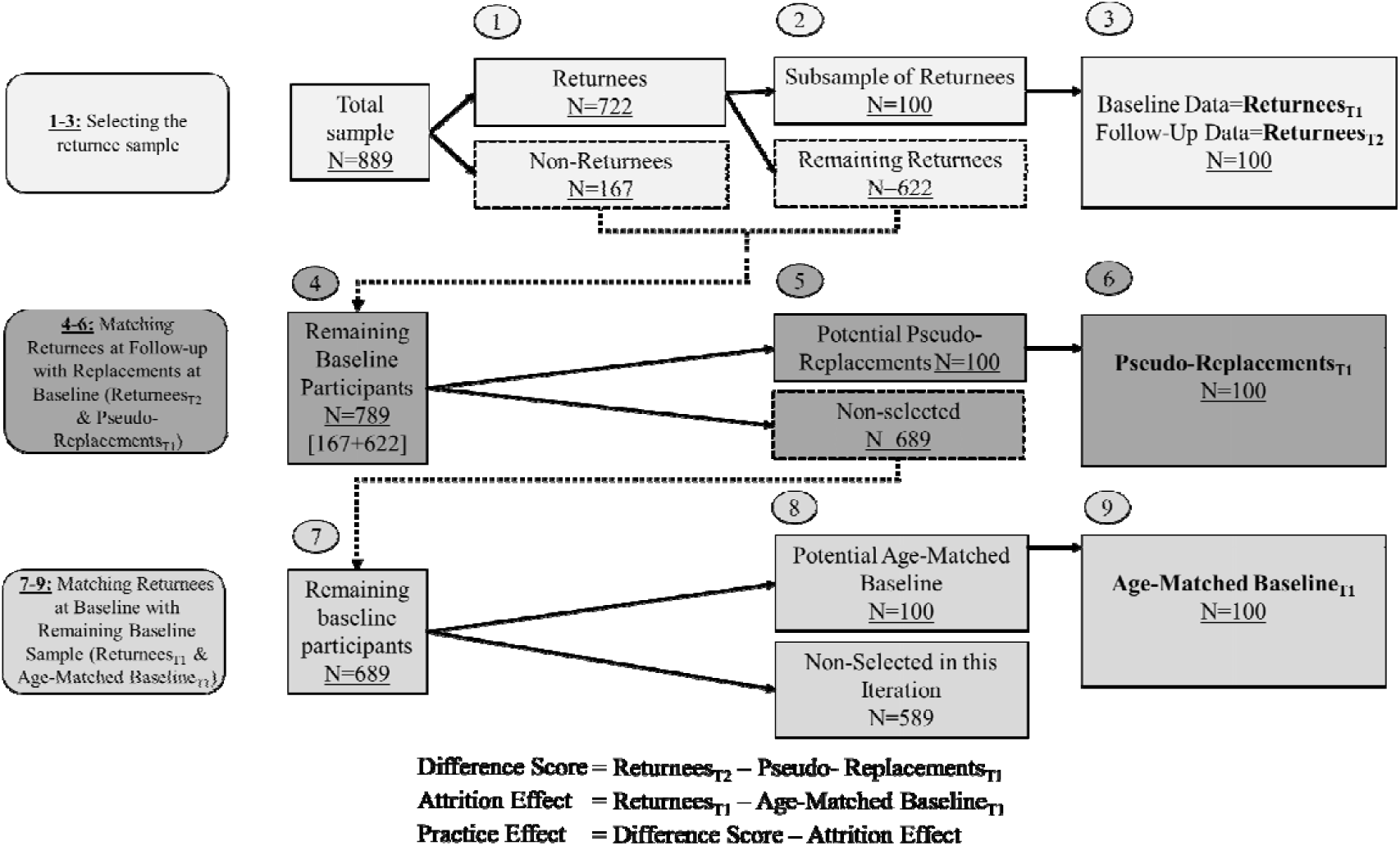
Participant selection and practice effect calculation within a single iteration of the pseudo-replacement method. Practice effects were calculated by comparing the mean scores of these subsamples using the equations provided below the flowchart. The difference between the mean of Returnees_T2_ scores and the mean of the matched Pseudo-Replacements_T1_ scores equates to the sum of practice effect and attrition effect. The attrition effect accounts for the fact that individuals who return for follow-up may be a higher performing or healthier than the full baseline sample. The practice effect for each test equals the difference score minus the attrition effect.

Practice effects were calculated by comparing the mean scores of these subsamples at each bootstrapping iteration as follows:

Difference score = Returnees_T2_ – Pseudo-Replacements_T1_

Attrition effect = Returnees_T1_ – Age-Matched Baseline_T1_

Practice effect = Difference score – Attrition Effect

The difference score represents the sum of the practice effect and the attrition effect. The attrition effect accounts for the fact that individuals who return for follow-up may be, on average, higher-performing or healthier than those who drop out. Since it is not possible to obtain an estimate of the mean outcome of the full baseline sample at the follow-up visit, we estimated an attrition effect by comparing returnees at baseline to the overall baseline sample for participants of a similar age at baseline. The bootstrapped nature of this approach results in an attrition effect that is matched at each iteration rather than an attrition effect based on those who truly attritted in ADNI. This improves the accuracy of each PE calculation because an individual iteration may be biased if the randomly selected returnees’ scores (Returnees_T1_) are significantly higher or lower than those of the age-matched baseline sample. Additionally, the age-matched baseline is randomly chosen and averaged across the entire bootstrapped analysis. This subsample retains the same distribution of returnees and attritors as the full baseline sample. Finally, the PE for each test equals the difference score minus the attrition effect.^10,16^ The PE for each test was then subtracted from each individual’s observed (unadjusted) follow-up test score to provide PE-adjusted raw scores.

In summary, this method identifies a comparison sample (pseudo-replacements) who are similar in age and other demographic characteristics to the returnees. The only difference between these samples is that returnees have taken the test before and pseudo-replacements have not. Because this analysis uses completed data, it requires no new participant recruitment, allowing the application of a replacement method of PE-adjustment to already completed studies. It is important to note that each participant’s baseline score could be placed in the matched baseline, the returnee baseline (if they also returned for the follow-up), or the pseudo-replacement sample depending on the bootstrap iteration. However, if that score was included in the pseudo-replacement sample, the participant was removed from the returnee sample and the matched-baseline sample for that iteration.

The samples are restricted at each iteration (approximately 100 pseudo replacements, 100 returnees, and 100 matched-baseline participants) in order to provide highly matched-groups. This matching preserves properties of statistical methods so they can be applied to statistically evaluate simulation results. All statistical methods require independence among observations or clusters of observations (such as baseline and follow-up data for each subject). If independence is severely compromised, for example, by using a larger sampling rate, corrections of asymptotic variance become necessary.^30^ As we are primarily interested in demonstrating presence of PEs (direction and magnitude) and accuracy of such evidence depends only on number of bootstrap replications, we used a reasonable group size (n = 100) to ensure stable estimates of PEs at each bootstrap iteration and applications of asymptotic variance without correction. The bootstrapped nature of this method allowed for a normal distribution of PEs.

Adjusted raw scores at follow-up were converted to z-scores, which were used to determine PE-adjusted diagnoses. In other words, determination of whether an individual was below the impairment threshold was now based on the PE-adjusted scores. McNemar χ^2^ tests were used to compare differences in the proportion of individuals classified as having MCI before and after adjusting for PEs, and to determine if PE-adjusted diagnoses increased the number of biomarker-negative CN participants and biomarker-positive MCI participants. Cohen’s d was calculated for each PE by comparing unadjusted and adjusted scores.

To determine the impact of PE adjustment in a clinical trial, we calculated sample size requirements for a hypothetical clinical trial aimed at reducing progression to MCI at 1-year follow-up in amyloid-positive CN individuals using MCI incidence rates from the present study. We performed logistic regressions with drug/placebo as the predictor and diagnosis at follow-up as the outcome. Sample size estimates were determined across a range of drug effects (10%-40% reduction in MCI diagnoses) with α=.05 and power=.80. We then used this information to estimate the effects on required sample size and cost for a variant of the Anti-Amyloid Treatment in Asymptomatic Alzheimer’s Disease (A4) Study given α=.05, and power=.80. The A4 Study recruited amyloid-positive CN individuals to investigate whether anti-amyloid therapy can delay cognitive decline.^3^ Progression to disease is a common and meaningful outcome for clinical trials. For our hypothetical variation of the A4 Study, the outcome of interest was progression to MCI at 1 year rather than just comparing cognitive decline. These analyses were completed within the powerMediation package in R v3.6.1.^31,32^

## RESULTS

PE magnitudes varied within and between cognitive domains (Table 1). PE-adjusted scores resulted in 113 (16%) converters to MCI; unadjusted scores resulted in 90 (12%) converters (Table 2A). Thus, there were 26% (p<.001) more individuals diagnosed with MCI after one year when using PE-adjusted scores (Table 2A). Table 2B shows that adjusting for PEs significantly increased the number of biomarker-positive participants who progressed to MCI (+15% to +20%) and decreased the number of biomarker-positive participants who remained CN (−5% to -6%). In particular, there was a 20% increase in amyloid-positive MCI cases and a 6% decrease in amyloid-positive CNs. These biomarker results suggest that PE-adjustment improved diagnostic accuracy. Table 3 shows the results when diagnoses were based on Petersen criteria. As can be seen in Table 3, although the proportions changed slightly, the pattern is the same as it was for the Jak-Bondi criteria. Moreover, all significant differences remained significant regardless of which diagnostic approach was implemented.

**Table 1:** Descriptive statistics and calculated practice effects for tests among participants who were cognitively normal at baseline

| Raw mean score<br>(SD) | <u>Memory</u> |  | <u>Attention/Executive</u><br><u>Function</u> |  | <u>Language</u> |  |
| --- | --- | --- | --- | --- | --- | --- |
|  | RAVLT | Logical<br>Memory | Trails A | Trails B | Boston<br>Naming | Category<br>Fluency |
| **All**<br>**baseline** | 8.25<br>(3.89) | 10.74<br>(4.28) | 34.57<br>(10.76) | 87.49<br>(40.27) | 27.49<br>(2.21) | 20.05<br>(5.25) |
| **Returnees Baseline** | 8.19<br>(3.69) | 10.53<br>(4.22) | 34.43<br>(10.87) | 86.21<br>(38.51) | 27.46<br>(2.14) | 20.08<br>(5.28) |
| **Returnees**<br>**Follow-Up** | 8.43<br>(4.47) | 11.65<br>(4.83) | 25.43<br>(12.63) | 86.22<br>(43.73) | 28.02<br>(2.13) | 20.17<br>(5.26) |
| **Replacements**<br>**Follow-Up,** | 8.16<br>(3.93) | 10.78<br>(4.37) | 34.91<br>(10.74) | 89.78<br>(42.77) | 27.46<br>(2.38) | 19.91<br>(5.20) |
| **Attrition**<br>**Effect** | -.06 | -.21 | -.17 | -.99 | -.03 | .03 |
| **Practice**<br>**Effect** | -.06 | 1.08 | -9.94 | -2.89 | .58 | .23 |
| **Cohen's**<br>**d** | .01 | .22 | .79 | .07 | .27 | .04 |

“All baseline” refers to all cognitively normal participants’ first assessment, including those that did not return for a 12-month visit. “Returnee Baseline” refers to baseline test scores for the portion of participants who returned for the 12-month follow-up visit (n=722). “Returnee Follow-Up” refers to 12-month scores for the portion of participants who returned for the 12-month follow-up (n=722).
“Replacement Follow-up” refers to the pseudo-replacement scores. The scores for memory tasks indicate the number of words remembered at the delayed recall trials. Scores on the attention/executive functioning tests indicate time to completion of task. On these tasks, higher scores indicate worse performance. Scores on the Boston Naming Task indicate number of correct items identified; scores on Category Fluency indicate number of items correctly stated. Practice effects and attrition effects are in raw units. As such, the negative practice effects and attrition effects for the Trails tasks demonstrates that practice decreased time (increased performance). Cohen’s d is given for the difference between PE-adjusted and unadjusted scores of returnees at follow-up. RAVLT= Rey Auditory Verbal Learning Test.

**Table 2.** Impact of practice effects

**A. Progression from cognitively normal to MCI**
| | # of cases,<br>based on<br>PE-unadjusted<br>cognitive scores | # of cases,<br>based on<br>PE-adjusted<br>cognitive scores | Difference<br>(%) # of cases | $\chi^2; p$ |
| --- | --- | --- | --- | --- |
| **MCI diagnosis** | 90 | 113 | +23 (+26%) | 20.0; <.001 |
| **Amnesic domain impaired** | 74 | 88 | +14 (+19%) | 12.1; .005 |
| **Attention/Executive**<br>**domain impaired** | 15 | 26 | +11 (+73%) | 9.01; .003 |
| **Language domain impaired** | 11 | 14 | +3 (+27%) | 1.3; .25 |

**B. Concordance of MCI diagnosis and biomarker-positivity**
| **Converters to MCI** | # of returnees<br>who are<br>biomarker-positive<br>and MCI<br>(PE-unadjusted) | # of returnees<br>who are<br>biomarker-positive<br>and MCI<br>(PE-adjusted) | Difference<br>(%) # of cases | *p* |
| --- | --- | --- | --- | --- |
| **Aβ+** | 44 | 53 | +9 (+20%) | .007 |
| **p-tau+** | 47 | 56 | +9 (+19%) | .031 |
| **t-tau+** | 40 | 46 | +6 (+15%) | <.001 |
| **Stable CN** | # of returnees who<br>are biomarker-positive and CN<br>(PE-unadjusted) | # of returnees who<br>are biomarker-positive and CN<br>(PE-adjusted) | Difference<br>(%) # of cases | *p* |
| **Aβ+** | 157 | 148 | -9 (-6%) | .007 |
| **p-tau+** | 175 | 166 | -9 (-5%) | .031 |
| **t-tau+** | 122 | 116 | -6 (-5%) | <.001 |

Follow-up diagnoses were made with practice effect-unadjusted (PE-unadjusted) or practice effect-adjusted (PE-adjusted) scores. The difference in the number of cases is calculated by subtracting the number of cases, based on PE-unadjusted scores, from the number of cases based on PE-adjusted scores. The percent difference (%) in number of cases is the differences in number of cases divided by the number of cases based on PE-unadjusted cognitive scores (e.g., 26%=23/90). $\chi^2$ is McNemar $\chi^2$ . Individuals could be impaired in more than one domain. Consequently, the sum of impaired individuals within each domain is greater than the total number of MCI cases. The MCI diagnosis row counts an individual only once, even if they are impaired in more than one domain.

Next, we showed that the number of participants necessary to determine significant drug effect across all effect sizes was substantially reduced when accounting for PEs (Figure 3). This is likely due to the increased base rate and more accurate diagnoses of PE-adjusted incident MCI. On average, adjusting for PEs reduced the number of participants required by 21.1% (533 participants) across effect sizes (range=114-2291 participants). The inset within Figure 3 focuses on differences for hypothetical PE-adjusted and unadjusted samples of ∼1,000.

**Figure 3:**
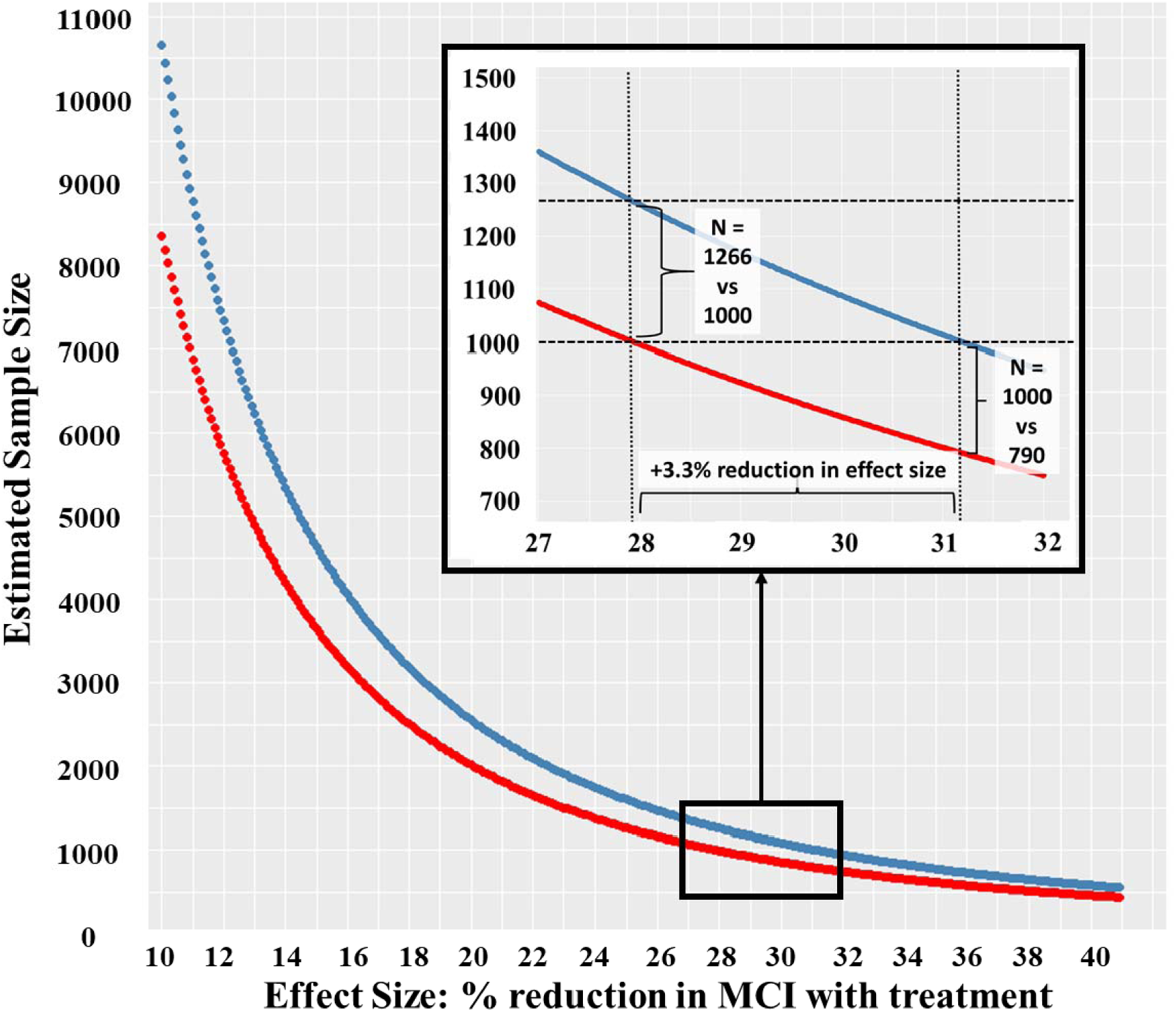
Effect of practice effect-adjusted vs unadjusted scores on a hypothetical clinical trial. Comparison of estimated sample sizes (Y-axis) necessary for detecting a significant drug effect (X-axis). The drug effect is operationalized as percent reduction in mild cognitive impairment (MCI) diagnoses at a 1-year follow-up between the treatment group and the placebo group. For example, a drug effect of 30% means that 30% more participants remained cognitively normal when treated with the drug than when given the placebo. The red line represents a trial that uses MCI incidence rates based on practice effect (PE)-adjusted diagnoses and the blue line represents a trial that uses incidence rates based on unadjusted diagnoses. MCI incidence rates were based on the subsample of participants from the present study who were amyloid-positive and cognitively normal at baseline. The model examined was a logistic regression with diagnosis at follow-up (MCI vs cognitively normal) as the outcome variable. The predictor was a two-level categorical variable representing placebo or drug. Alpha was set at .05, power was .80, and the hypothetical sample was evenly split into treatment and placebo groups. Across all effect sizes (10%-40% reduction in treatment vs placebo conversation rates) the PE-adjusted trial required fewer participants than the PE-unadjusted trial. The inset shows results for hypothetical samples with ∼1000 participants. If this study used PE-unadjusted outcome measures (blue line), it would require n effect size of 31.2% to reach a significant result with ∼1000 participants. Using PE-adjusted diagnoses, only 790 participants would be required for the same study with the same drug effect, a reduction of 210 participants. A PE-adjusted study with ∼1000 participants (red line in the inset) would be able to detect a smaller drug effect of 27.9%. With this 3.3% reduction in effect size, a PE-unadjusted study would require an additional 266 participants at this drug effect level (1266 vs 1000).

We then applied our findings to recruitment data from the A4 Study^33^ (Figure 4A). Obtaining the cognitively normal, amyloid positive A4 sample of 1323 required the recruitment of 5.11 times as many people for initial screening (n=6763) and 3.39 times as many people to undergo amyloid PET imaging (4486).^33^ Our calculations showed that this sample size of 1323 would be powered to detect a 27.4% drug effect on incident MCI outcomes. Due to the higher base rates of incident MCI at follow-up after PE adjustment, calculations from the R powerMediation package showed that only 1045 participants would be required. As shown in Figure 4A, the number of initial screens and amyloid PET scans would, in turn, be substantially reduced to 5340 and 3543, respectively. Figure 4B also shows the range of sample size reductions for differing drug effect sizes for initial screening (reduced ns from 583 to 11711) and amyloid PET imaging (reduced ns from 386 to7768). As estimated drug effect sizes gets smaller, the reductions in necessary sample size, and the cost reductions that those reductions would engender, get substantially larger.

**Figure 4:**
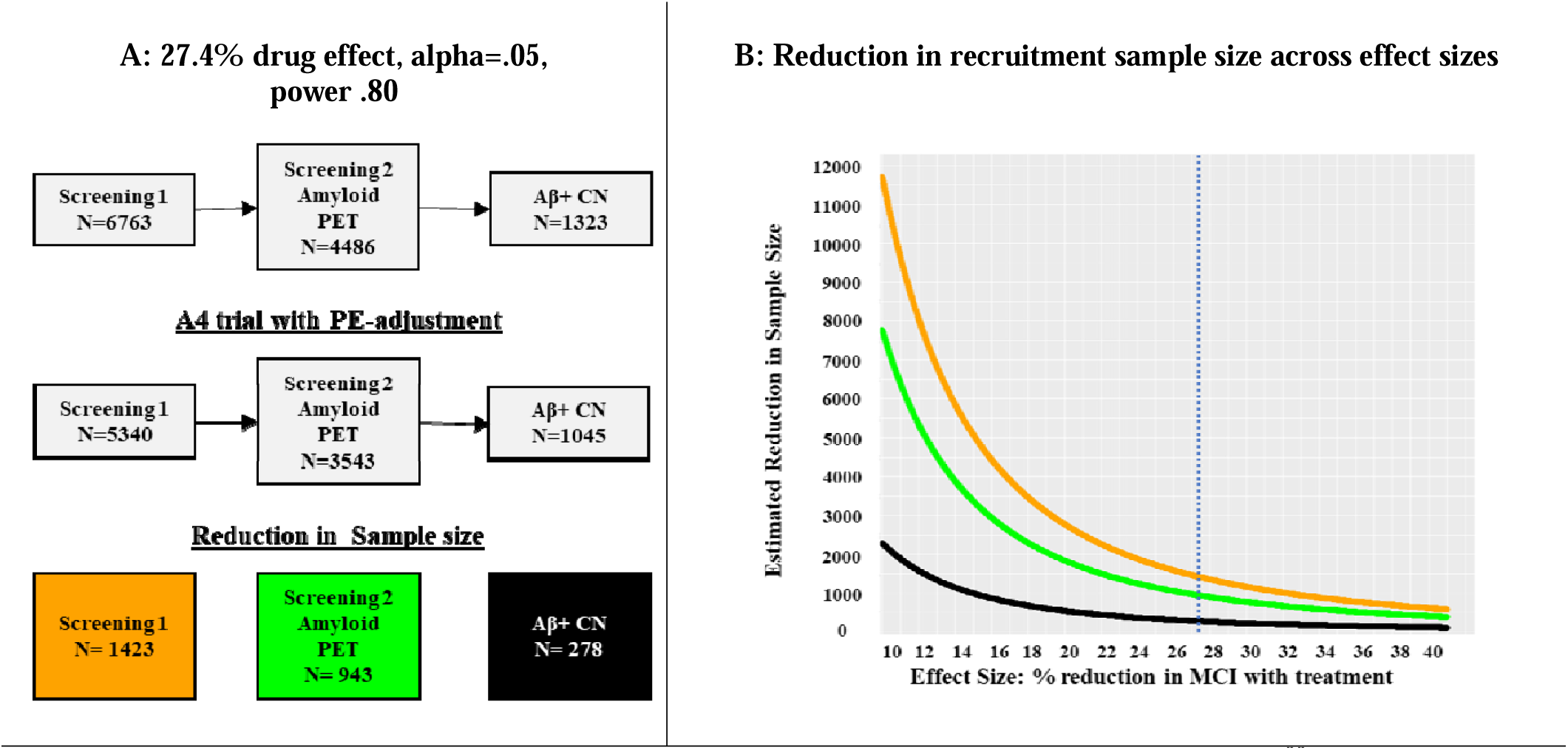
Comparison of recruitment design for detection of a drug effect based on A4 Study^33^ recruitment. Using sample size estimates from Figure 3, we present how planning to adjust for practice effects would alter a clinical drug trial, using A4 Study recruitment as an example. The A4 Study had a total sample of 1323 participants after recruitment as shown in the top row of gray boxes (based on Figure 1 in Sperling et al., 2020). [25] A: Based on sample size estimates from Figure 3, a sample of 1323 would enable a study to detect a significant drug effect of 27.4% at an alpha of .05 and .80 power. The top row of the flow chart presents the recruitment for the A4 study. This study reported an initial screening (6763 participants) followed by amyloid PET (4486 participants) imaging to achieve their sample of 1323 amyloid-positive (AB+), cognitively normal (CN) participants. Achieving the final sample size thus required an n for the initial screening that was 5.11 times as large as the final sample size, and an n for amyloid PET imaging that was 3.39 times as large as the final sample. Our power analyses suggest that the same effect size is achieved with only 1042 participants if a trial adjusted follow-up scores for practice effects. That, along with the reductions in initial screening and PET scans, is shown in the middle row of the flow chart. The bottom row shows the sample size reductions for initial screening, PET screening, and the initial biomarker-positive and cognitively normal sample. **B**: The figure presents the reduction in recruitment sample size (Y-axis) across effect sizes ranging from 10% to 40% (X-axis). The orange line represents how many fewer participants would be necessary at initial screening if a study had planned to adjust for practice effects at follow-up. The green line represents how many fewer participants would be necessary for amyloid PET imaging if a study had planned to adjust for practice effects at follow-up. The black line represents what the final sample size would be. The numbers were calculated for each effect size in the way described in panel A. Each curve was calculated for each effect size at 1% increments in the manner described in panel A. The dotted blue line shows at effect size of 27.4%.

## DISCUSSION

Accounting for cognitive PEs resulted in a 26% increase in 12-month MCI incidence. The increase in biomarker-positive MCI (+20% amyloid-positive) and reduction in biomarker-positive CN participants (−6% amyloid-positive) supports diagnostic validity. Therefore, this approach reduces the observed discrepancy between biologically- and clinically-based diagnoses.^34^ Importantly, given 26% more cases in only 12 months, these results confirm our hypothesis that adjusting for PEs using this method leads to earlier detection of MCI. Individuals diagnosed with MCI based on PE-adjusted scores—who would otherwise have been considered CN—would be expected to progress to AD dementia sooner than true CN participants. Progression at later follow-ups (24 months, 36 months) was consistent with this hypothesis, but sample sizes were too small for statistical comparisons.

Results of the present study suggest that failure to account for PEs leads to a substantial number of false negatives as 18% of biomarker-positive MCI cases were labeled as CN at follow-up. Accounting for PEs also appeared to improve accuracy, reducing false positives by 5%. As such, not adjusting for PEs will weaken our ability to accurately determine the effect of novel treatments and to compare biomarker differences between cases and controls, a goal of current research guidelines.^35^

To quantify how clinical trials would be improved by PE adjustment, we estimated sample sizes necessary to power a simulated clinical drug trial. Other things being equal, detecting differences or making predictions is less accurate for low base rate events,^36^ and our PE adjustment increased the base rate of MCI at 12-month follow-up. Progression to disease is the most common outcome of interest in clinical trials, and results indicate that smaller samples would be needed for clinical trials with a PE-adjusted endpoint. Across effect sizes, there was an average reduction of 21% in necessary sample size when using PE-adjusted diagnoses; sample size reductions were greater with smaller treatment effect sizes (Figures 3, 4a). Based on screening/recruitment numbers in the A4 Study,^33^ Figure 4A showed that determining conversion to MCI using PE-adjusted scores would mean 1423 fewer initial screenings and 943 fewer amyloid PET scans. At $5000 per scan, cost savings for that alone would be $4.72 million. Initial screening—which included cognitive testing, clinical assessments, and *APOE* genotyping—for 1423 individuals would result in considerable additional cost savings, estimated at $3.35 million. Cost saving will be partially offset by needing additional participants to serve as replacement participants for calculating PEs. In prior work, 150-200 replacement participants was sufficient.^10^ With replacements for 3 follow-up sessions with 200 participants each, we estimated additional costs of $615,000. Estimated overall savings would be $7.45 million. Moreover, PE-adjusted diagnoses result in earlier detection, which mean shorter follow-up periods. Reduced study duration would lead to still further cost reductions and benefits including lower participant and staff burden, fewer invasive procedures, and likely reduced attrition.

Delayed detection of MCI is extremely costly from a public health perspective. In 2018, the Alzheimer’s Association projected an estimated U.S. national savings of $231 billion by 2050 if those on the AD-trajectory were diagnosed during the MCI, rather than the dementia, stage.^1^ In clinical practice, the MCI stage represents a critical time for preparation and intervention for individuals who will progress to AD-related dementia. If PEs delay detection of MCI, clinicians may also be providing inadequate care to those most at risk.

A 2012 meta-analysis and 2015 review described several approaches to estimating PEs.^9,13^ One method is not necessarily better than another, and some simply address different issues. Almost all non-replacement approaches are only informative about relative differences or focus on change scores rather than producing stand-alone follow-up scores adjusted for PEs. One approach is to retest participants after a short interval and use the second testing as the comparison for future assessments. Individuals with smaller PEs at 1 week are more likely to have worse baseline biomarker profiles and experience steeper 1-year decline compared to other participants.^14,28,29^ However, this method requires an additional test session for the entire sample. Other studies have found that additional baseline tests improves prediction of progression to MCI ^7,37-41^ Whether complete retesting after a week improves prediction over the less burdensome and less costly inclusion of additional measures at baseline testing remains to be determined. Similarly, regression-based methods require a large, normative change sample. Change is assessed relative to that sample, but PEs are still unknown in the normative sample. Both methods gauge only relative change. They cannot be used for absolute diagnostic cutoff thresholds, and thus cannot have any effect on when a person crosses an impairment threshold resulting in conversion to MCI. Nor can they calculate PEs in the presence of a mean-level decline over time, which is expected in older adults. The replacement-participants method requires a small number of additional participants relative to an entire study sample. It generates adjusted scores at follow-up that are not obscured by age-related decline. The other methods can compare trajectories of people already diagnosed as MCI or CN, but only the replacement method—which generates absolute PE-adjusted scores—can alter when MCI is detected. Although the replacement-participants method reduces all scores, it does not change individual differences in any way. Thus, it also allows for comparison of trajectories.

Surprisingly, we found no practice effect on the AVLT. This may have occurred because, despite receiving the same version at the 12-month visit, some participants also completed an alternate version of the AVLT at a 6-month visit. The reduced 12-month practice effect for AVLT is consistent with the well-known phenomenon of retroactive interference, i.e., the different 6-month version interfering with the PE from exposure to the baseline/follow-up version. Prior studies, including our own, have consistently found PEs on the AVLT or similar episodic memory measures.^9,10,16^ Thus, the present estimate of the impact of PEs may be a conservative one. It is also noteworthy that despite the lack of an apparent AVLT practice effect in the current study, we still found an increase in amnestic MCI cases after adjusting for PEs. This highlights the importance of including more than one test in each cognitive domain as specified in the Jak-Bondi approach.^7,8,10,37^ Finally, we note that use of alternate forms is considered suboptimal as even well-matched forms are not equivalent and add an additional source of test-retest variance.^42^

The replacement-participant method is not dependent on any particular MCI diagnosis approach. Percent changes differed for Jak-Bondi and Petersen criteria, but all significant differences remained for both diagnostic approaches.

We acknowledge some limitations of the study. ADNI is not a population-based study and is not representative of the general population in terms of sociodemographic factors. However, replacement methods have been shown to be effective in other studies, including population-based samples.^10,16^ The method currently only examines PEs across 2 time points. As PEs persist over time, their magnitude may differ with additional assessments. We do not know what the effects would be in cases with multiple follow-up visits. Future studies will benefit from PE adjustments that are able to separate age-related decline from PEs over multiple retest visits. As noted, including matched replacements for third and fourth visits could still be cost-effective. Some participants who do not qualify after initial screening or those who do not agree to biomarker assessment might still qualify to serve as replacement participants. Ultimately, the field would benefit from the development of PE norms, particularly those stratified by age and testing interval, which could obviate the need for replacement participants. Importantly, the PE magnitudes in these analyses should not be directly used in any other study. Our results and those of other studies suggest that PEs are often sample specific and that the need to be calculated for each study.^9,27^ The above sample size estimations are a hypothetical example of how the method may affect a clinical trial. The goal was to provide more empirical evidence supporting the use of PE-adjustment, as suggested by many other studies.^13,14^

In summary, adjusting for PEs results in earlier and more accurate detection of MCI. Reluctance to include additional replacement-participant testing is understandable as it increases cost and participant burden. In the end, however, it would substantially reduce the necessary sample size, follow-up time, burden, and cost for clinical trials or other longitudinal studies. Although the magnitude of PEs may not be generalized from one sample to another, the method is appropriate for all ages and retest intervals because replacements are always matched on these features. The method is also not dependent on any specific approach to the diagnosis of MCI. Additionally, have shown that the replacement-participant method can be adapted for ongoing or already completed studies that did not specifically recruit matched-replacement participants. Given the public health importance of the earliest possible identification of AD pathology, it is strongly recommended that accounting for PEs be a planned component of clinical trials, routine clinical work, and longitudinal studies of aging and aging-related cognitive disorders.

## Data Availability

The data used by this project are from the Alzheimer's Disease Neuroimaging Initiative (ADNI). For access to this data, please see their data use agreement procedures: http://adni.loni.usc.edu/data-samples/access-data/

## Acknowledgments

The content of this article is the responsibility of the authors and does not necessarily represent official views of the National institute of Aging or the Department of Veterans affairs. The ADNI and funding sources had no role in data analysis, interpretation, or writing of this project. The corresponding author was granted access to the data by ADNI and conducted the analyses. The study was supported by grants from the U.S. National institute on Aging (MSC: F31AG064834, WSK, CEF, MJL: R01 AG050595, CEF, WSK: P01 AG055367; WSK, R01 AG022381, AG054002, AG060470; CEF, R01 AG059329; ALG: K01 AG050699; MWB: R01 AG049810) and the National Center for Advancing Translational Sciences (JAE: KL2 TR001444). The Center for Stress and Mental Health in the Veterans Affairs San Diego Healthcare System also provided support for this study.

## Author contributors

The study was conceived by MSC and WSK. Guidance on statistical analysis was provided by XMT and ALG. Determination of MCI diagnoses was made by ECE, MWB, JE, KRT. MSC, WSK, JAE, MSP, and DEG contributed to the practice effects methodology. Primary funding to support this work was obtain by WSK, CEF, MJL, and MSC. All authors provided critical review and commentary on the manuscript.

